# The impact of work loss on mental and physical health during the COVID-19 pandemic: Findings from a prospective cohort study

**DOI:** 10.1101/2020.09.06.20189514

**Authors:** Daniel Griffiths, Luke Sheehan, Caryn van Vreden, Dennis Petrie, Genevieve Grant, Peter Whiteford, Malcolm R Sim, Alex Collie

## Abstract

**Objective:** To determine if losing work during the COVID-19 pandemic is associated with mental and physical health. To determine if social interactions and financial resources moderate the relationship between work loss and health.

**Design:** Baseline data from a prospective longitudinal cohort study.

**Setting:** Australia, 27^th^ March to 12^th^ June 2020.

**Participants:** Australians aged 18+ years, employed in a paid job prior to the COVID-19 pandemic who responded to an online or telephone survey.

**Main Outcome Measures:** Kessler-6 score > 18 indicating high psychological distress. Short Form 12 (SF-12) mental health or physical health component score < = 45 indicating poor mental or physical health.

**Results:** 2,603 respondents including groups who had lost their job (N = 541), were not working but remained employed (N = 613), were working less (N = 789) and whose work was unaffected (N = 789). Three groups experiencing work loss had greater odds of high psychological distress (AOR = 2.22–3.66), poor mental (AOR = 1.78–2.27) and physical health (AOR = 2.10–2.12) than the unaffected work group. Poor mental health was more common than poor physical health. The odds of high psychological distress (AOR = 5.43–8.36), poor mental (AOR = 1.92–4.53) and physical health (AOR = 1.93–3.90) were increased in those reporting fewer social interactions or less financial resources.

**Conclusion:** Losing work during the COVID-19 pandemic is associated with mental and physical health problems, and this relationship is moderated by social interactions and financial resources. Responses that increase financial security and enhance social connections may partially alleviate the health impacts of work loss.

**Registration:** Australian New Zealand Clinical Trials Registry: ACTRN12620000857909.

## Introduction

Changes in work, including widescale unemployment and reductions in working hours, have been one of the major consequences of public health measures taken to limit transmission of the Severe Acute Respiratory Syndrome Coronavirus 2 (SARS-CoV-2), which leads to the Coronavirus Disease 2019 (COVID-19). Globally there was an estimated 14% reduction in working hours during the first half of 2020 compared to the last quarter of 2019, equivalent to a loss of 400 million full-time jobs (1). In Australia, an estimated 870,000 workers (6.7% of the total employed) lost their jobs between February and May 2020 after widespread public health measures to contain COVID-19 were introduced, while in May a further 1.55 million remained employed but were working less or were stood down (2). A coronavirus-induced recession with widespread job losses has potential to lead to an epidemic of mental illness, chronic disease and mortality (3). Australian data demonstrates a higher than normal prevalence of stress, anxiety and hopelessness among the general community during the COVID-19 pandemic (4), and elevations in the community prevalence of depression and anxiety symptoms (5).

Work and health are closely interconnected. There is substantial evidence globally of the health benefits of good work (6). The harmful health impacts of losing work are also well described (7), including in people whose work is impacted by viral epidemics (8, 9). Work loss both disrupts social connections and reduces material financial resources, which are important determinants of health (10, 11).

The response to the employment crisis by the Australian Government has been temporary wage subsidies (12), and increases in social security payments (13), to ensure workers maintain employer connections and a source of income. The ability of Australian workers to maintain social interaction has been challenged by physical distancing, isolation, movement restriction and working from home requirements. The health of those that have maintained higher levels of social interaction, and who have greater financial resources, may be less impacted by work loss. This study aimed to determine whether losing work during the COVID-19 pandemic is associated with poorer mental and physical health, and to determine if financial resources and social interactions moderate the relationship between work loss and health. We hypothesised that a gradient in work loss would be reflected in a health gradient, with the most affected group (the newly unemployed) reporting the worst health status.

## Methods

### Design, setting and participants

We report findings from a baseline survey of a prospective longitudinal cohort study of people living in Australia, aged at least 18 years, and who were employed in a paid job or self-employed prior to the COVID-19 pandemic.

Participants completed a 20-minute baseline survey upon enrolment (either online or via a telephone survey) between 27^th^ March and 12^th^ June 2020, which included both standardised health metrics and a range of study-specific questions. The online survey was promoted via social and general media, through personal networks, and via newsletters distributed by community sector and industry groups. Participants were enrolled into the telephone survey via random digit dialling conducted by a third-party market research company.

Four study groups were defined on the basis of changes in work and job loss at the time of the baseline survey: (i) *Lost Job* – those who had lost their jobs during the COVID-19 pandemic and were not working, (ii) *Off-Work* – those still employed but not currently working (e.g. stood down, furloughed, taking leave), (iii) *Reduced Work* – those still employed and working fewer hours than before the pandemic (e.g. reduced fraction, fewer days working), and (iv) *Work Unaffected* – those employed and working the same or more hours as before the pandemic.

### Health outcomes

Psychological distress was assessed using the Kessler 6 scale (14), distinguishing levels of serious mental illness (15). Mental and physical health were assessed using the Mental Component Score and the Physical Component Score of the Short Form 12 (SF-12) scale (16). Outcome measures were derived using standardised approaches and included whether the respondent recorded:

- A Kessler-6 score of more than 18 indicating high psychological distress
- A SF-12 mental health component score of less than 45 indicating poor mental health.
- A SF-12 physical health component score of less than 45 indicating poor physical health.

### Social interactions, financial resources and demographics

Financial resources were assessed with the question: ‘If all of a sudden you had to get $2000 for something important, could the money be obtained within a week?’ (17). Responses of ‘yes’ were categorised as having more financial resources and responses of ‘no’ and ‘don’t know’ as having less financial resource. Social interactions were measured using the Social Interaction sub-scale of the Duke Social Support Index (18). Scores were dichotomised with reference to the cohort median as (1) less social interaction for scores less than 8, or (2) high levels of social interaction for scores greater than or equal to 8.

Other survey items included a range of sociodemographic, work history and health characteristics including age, gender, household circumstances, personal income, pre-COVID-19 occupation and working hours, and the presence of pre-existing medical conditions.

### Analytical approach

The work-health relationship was modelled using binary logistic regression with working status group as the independent variable. Models included interaction terms between working status with social interaction, and working status with financial resources, in addition to adjusting for age, gender, preexisting medical conditions and survey mode. The reference group for interaction terms was the Unaffected Work subgroup with more social interactions and more financial resources. Pre-existing diagnosed anxiety and depression were included as variables in models for psychological distress and mental health. For the physical health model, a diverse set of health conditions were compiled into the total number of medical conditions variable. Preliminary analysis identified differences in cohort profiles and outcomes between survey modes and thus a variable for survey mode was also included.

## Results

A total of 2603 participants enrolled in the study, including 541 (20.8%) in the Lost Job group, 613 (23.5%) in the Off-Work group, 660 (25.4%) in the Reduced Work group and 789 (30.3%) in the Unaffected Work group. For each group, the proportion completing the survey via telephone was 23.8% (Lost Job), 20.4% (Off-Work), 51.1% (Reduced Work) and 100% (Unaffected Work).

### Psychological distress

Across the cohort 438 respondents (17.1%) reported high psychological distress, and its prevalence differed by working status (Table 1). A total of 159 (30.0%) respondents from the Lost Job group demonstrated high psychological distress compared with 156 (25.9%) from the Off-Work group, 108(16.7%) from the Reduced Work group, and 15 (1.9%) from the Unaffected Work group.

**Table 1.** Summary statistics for health outcomes and predictor variables.

|  | Whole cohort<br>N (col%) | Psychological distress |  | Mental health |  | Physical health |  |
| --- | --- | --- | --- | --- | --- | --- | --- |
|  |  | High | Low or moderate | Poor | Average or good | Poor | Average or good |
|  | 2603<br>(100.0%) | 438<br>(17.1%) | 2119<br>(82.9%) | 1348<br>(51.9%) | 1249<br>(48.1%) | 315<br>(12.1%) | 2281<br>(87.9%) |
| **Working Status** |  |  |  |  |  |  |  |
| Lost Job | 541<br>(20.8%) | 159<br>(30.0%) | 371<br>(70.0%) | 374<br>(69.1%) | 167<br>(30.9%) | 75<br>(13.9%) | 466<br>(86.1%) |
| Off-Work | 613<br>(23.5%) | 156<br>(25.9%) | 446<br>(74.1%) | 392<br>(64.1%) | 220<br>(35.9%) | 82<br>(13.4%) | 530<br>(86.6%) |
| Reduced Work | 660<br>(25.4%) | 108<br>(16.7%) | 539<br>(83.3%) | 392<br>(59.5%) | 267<br>(40.5%) | 92<br>(14.0%) | 566<br>(86.0%) |
| Unaffected Work | 789<br>(30.3%) | 15<br>(1.9%) | 763<br>(98.1%) | 190<br>(24.2%) | 595<br>(75.8%) | 66<br>(8.4%) | 719<br>(91.6%) |
| **Social interaction** |  |  |  |  |  |  |  |
| Less | 1393<br>(53.5%) | 308<br>(27.4%) | 817<br>(72.6%) | 736<br>(65.1%) | 395<br>(34.9%) | 165<br>(14.6%) | 966<br>(85.4%) |
| More | 1134<br>(43.6%) | 114<br>(8.3%) | 1263<br>(91.7%) | 554<br>(39.9%) | 836<br>(60.1%) | 134<br>(9.6%) | 1255<br>(90.4%) |
| **Financial resource** |  |  |  |  |  |  |  |
| Less | 763<br>(29.3%) | 269<br>(35.5%) | 489<br>(64.5%) | 563<br>(74.0%) | 198<br>(26.0%) | 126<br>(16.6%) | 634<br>(83.4%) |
| More | 1805<br>(69.3%) | 168<br>(9.4%) | 1618<br>(90.6%) | 766<br>(42.5%) | 1035<br>(57.5%) | 180<br>(10.0%) | 1621<br>(90.0%) |
| **Gender** |  |  |  |  |  |  |  |
| Female | 1614<br>(62.0%) | 337<br>(21.2%) | 1253<br>(78.8%) | 948<br>(58.8%) | 665<br>(41.2%) | 201<br>(12.5%) | 1411<br>(87.5%) |
| Male | 975<br>(37.5%) | 96<br>(10.1%) | 858<br>(89.9%) | 394<br>(40.6%) | 576<br>(59.4%) | 112<br>(11.5%) | 858<br>(88.5%) |
| **Age** |  |  |  |  |  |  |  |
| 18-24 years | 245<br>(9.4%) | 49<br>(20.4%) | 191<br>(79.6%) | 142<br>(58.2%) | 102<br>(41.8%) | 20<br>(8.2%) | 224<br>(91.8%) |
| 25-34 years | 442<br>(17.0%) | 87<br>(20.1%) | 345<br>(79.9%) | 259<br>(59.0%) | 180<br>(41.0%) | 37<br>(8.4%) | 402<br>(91.6%) |
| 35-44 years | 484<br>(18.6%) | 89<br>(18.8%) | 385<br>(81.2%) | 273<br>(56.5%) | 210<br>(43.5%) | 44<br>(9.1%) | 439<br>(90.9%) |
| 45-54 years | 647<br>(24.9%) | 121<br>(19.1%) | 514<br>(80.9%) | 352<br>(54.4%) | 295<br>(45.6%) | 76<br>(11.7%) | 571<br>(88.3%) |
| 55-65 years | 652<br>(25.0%) | 87<br>(13.5%) | 557<br>(86.5%) | 296<br>(45.5%) | 355<br>(54.5%) | 115<br>(17.7%) | 535<br>(82.3%) |
| 65+ years | 113<br>(5.1%) | 5<br>(3.8%) | 127<br>(96.2%) | 26<br>(19.5%) | 107<br>(80.5%) | 23<br>(17.3%) | 110<br>(82.7%) |
| **Pre-existing medical conditions** |  |  |  |  |  |  |  |
| Depression | 492<br>(18.9%) | 188<br>(38.3%) | 303<br>(61.7%) | 391<br>(79.5%) | 101<br>(20.5%) |  |  |
| Anxiety | 463<br>(17.8%) | 191<br>(41.3%) | 272<br>(58.7%) | 378<br>(81.6%) | 85<br>(18.4%) |  |  |
| Mean (SD) number | 1.0<br>(1.6) |  |  |  |  | 2.0<br>(2.4) | 0.9<br>(1.5) |
| **Survey mode** |  |  |  |  |  |  |  |
| Online | 1223<br>(47.0%) | 383<br>(31.9%) | 819<br>(68.1%) | 902<br>(73.8%) | 321<br>(26.2%) | 161<br>(13.2%) | 1062<br>(86.8%) |
| Telephone | 1380<br>(53.0%) | 55<br>(4.1%) | 1300<br>(95.9%) | 446<br>(32.5%) | 928<br>(67.5%) | 154<br>(11.2%) | 1219<br>(88.8%) |

Note: All data is reported as number (row percentage) unless otherwise stated. SD = standard deviation.

For subgroups with more social interaction and more financial resources, the adjusted odds of high psychological distress were significantly greater for the Lost Job group (AOR = 3.66 [1.47, 9.14]) and the Off-Work group (AOR = 3.35 [1.38, 8.16]) compared with the reference group, indicating a relationship between a gradient in exposure to work and psychological distress (Table 2). Lower levels of social interaction was associated with greater odds of high psychological distress across the three subgroups with work loss (AOR = 5.83–7.25). Less financial resources was associated with greater odds of high psychological distress across all groups (AOR = 5.43–8.36).

**Table 2.**
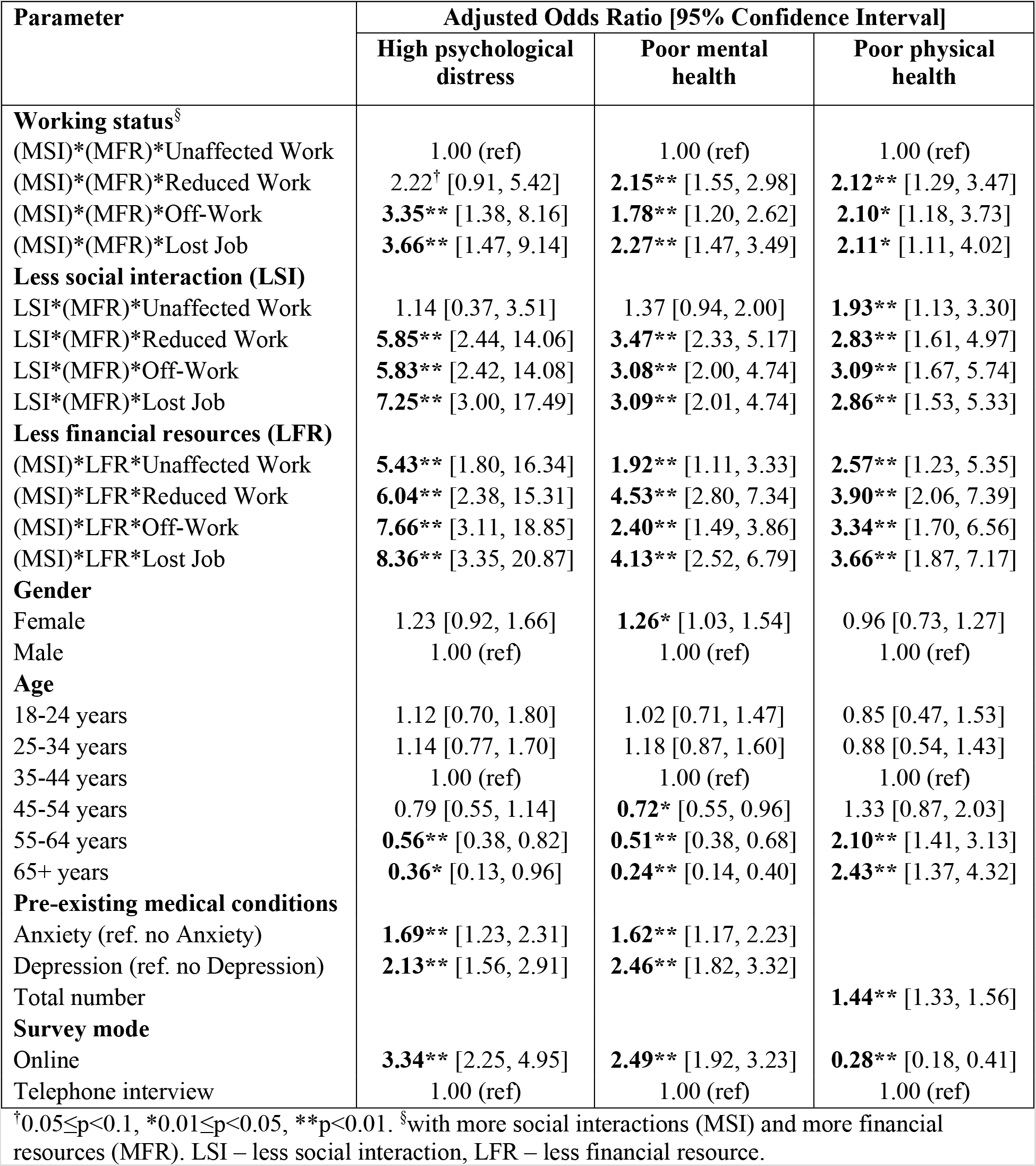
Binary logistic regression models for psychological distress, mental health and physical health outcomes.

### Mental health

Poor mental health was common in the cohort, occurring in 1348 (51.9%) of all respondents. Prevalence was lowest in the Unaffected Work group with 190 (24.2%) of respondents recording poor mental health, compared with 392 (59.5%) in the Reduced Work group, 392 (64.1%) in the Off-Work group, and 374 (69.1%) in the Lost Job group.

For respondents with more social interaction and more financial resources, the adjusted odds of poor mental health were significantly greater for the Reduced Work group (AOR = 2.15 [1.55, 2.98]), Off-Work group (AOR = 1.78 [1.20, 2.62]), and the Lost Job group (AOR = 2.27 [1.47, 3.49]) compared with the Unaffected Work group. Less social interaction was associated with greater odds of poor mental health for those in the three work loss groups (AOR = 3.08–3.47). Possessing less financial resources was associated with greater odds of poor mental health, across all working status groups (AOR = 1.92–4.53), including the Unaffected Work group.

### Physical health

The prevalence of poor physical health was much lower than for poor mental health across the cohort, constituting 315 (12.1%) respondents. The Unaffected Work group had 66 (8.4%) respondents with poor physical health, compared with 92 (14.0%) for the Reduced Work group, 82 (13.4%) for the Off-Work group, and 75 (13.9%) for the Lost Job group.

The odds of poor physical health, for those with a more social interaction and more financial resources, were significantly higher for those in the three work loss groups (AOR = 2.10–2.12) compared with the Unaffected Work group. Less social interaction was associated with greater odds of poor physical health across all working status groups (AOR = 1.93–3.09), as was less financial resources (AOR = 2.57–3.90).

### Covariates

A total of 1128 (43.4%) respondents reported having at least one pre-existing medical condition, including 463 (17.8%) with anxiety and 492 (18.9%) with depression (Table 1). Respondents who reported anxiety or depression had greater odds of high psychological distress and poor mental health. For physical health, each additional medical condition reported increased the odds of poor physical health by 1.44. Female respondents had greater odds of poor mental health than male respondents. Age-related differences were observed across all health outcomes. Older respondents had lower odds of psychological distress and poor mental health than the reference group of those aged 35 to 44 years old, but higher odds of poor physical health. Respondents who completed the survey online had greater odds of high psychological distress and poor mental health and lower odds of poor physical health compared to those completing the survey via telephone.

## Discussion

This study demonstrates that in a cohort of people employed prior to the COVID-19 pandemic, those experiencing work loss are more likely to report psychological distress, and poor mental and physical health compared to those whose work was unaffected. These negative health effects are exacerbated in people reporting fewer financial resources and those reporting lower levels of social interaction. These findings demonstrate that financial hardship and social connections moderate the relationship between work and health in the extraordinary circumstances of the COVID-19 pandemic (7). Strategies that promote social interactions and increase financial security in those experiencing job or work loss may help to minimise negative health impacts.

The odds of high psychological distress were greatest in people reporting lower financial resources. Among these respondents, the greatest odds of distress were reported by people who had lost their job, followed by those who were off work, those with reduced working hours and finally by those whose work was unaffected. The odds of poor mental and physical health were also greater in people reporting lower financial resources. The links between financial resources and health, particularly mental health, are well established (19). Our data additionally show that lower access to financial resources exacerbates the negative health consequences of work loss. People with lower financial reserves and more likely to report increases in financial stress (20). Potential interventions to ameliorate these impacts include increasing the financial support available to those who have lost work during the pandemic, and supporting people to manage their existing financial resources.

The Australian government introduced two major economic stimulus programs to provide financial support to people whose work was affected during the COVID-19 pandemic (12, 13). Our findings suggest that these economic measures may have had positive health consequences, by reducing the number of people in financial stress early during the pandemic. Conversely, withdrawal of these measures will reduce access to financial resources and may result in a worsening of mental and physical health among working age Australians. Enhancing access to services such as financial planning or financial counselling may also be helpful.

We also observed that social interactions moderated the work-health relationship. Social support is an important determinant of health. Loneliness and social isolation have been associated with increased mortality, as well as adverse physical and mental health outcomes (21). During the COVID-19 pandemic social interactions have been transformed by the public health measures introduced to reduce viral transmission. The ability to participate in activities that support health such as working or volunteering, meeting in groups, participating in clubs and sporting groups (22) have all been reduced. Importantly, we observe that the moderating impact of lower social isolations on mental health and psychological distress was limited to people in the Lost Job, Off Work and Reduced Work groups. Those whose work was unaffected and who reported low social interactions did not report elevated distress or poorer mental health than their counterparts with more social interactions, suggesting a protective effect of continued engagement in work.

Our data also provide some evidence for a gradient in health that can be related to the extent of work loss. Those in the job loss group had the greatest odds of reporting high psychological distress, poor mental and physical health than those in the other study groups. Those in the unaffected work group had the lowest odds of reporting these adverse health outcomes, while the reduced work and off work groups were intermediate.

Economic recovery from the pandemic is likely to be a long-term process, and is unlikely to be evenly spread across society. A second wave in community transmission in Victoria (23) has led to further job and work losses, more stringent movement restrictions, and business closures. Some industries and occupations are at greater risk of pandemic-linked work loss than others (24), due to the inability to work remotely or enforce physical distancing, the rate of insecure and casual work arrangements, the risk of workplace transmission to employees or members of the public, or being considered ‘nonessential’ and thus more susceptible to business closure during outbreaks. Health-promoting programs should be targeted to people working in these high-risk industries and occupations, and to those whose working arrangements mean they are ineligible for alternative forms of financial assistance.

To our knowledge, this is one of the first studies examining psychological distress, mental and physical health, specifically among people losing work during the COVID-19 pandemic. Study strengths include the use of validated measurement instruments, the temporal proximity of data collection to job and work loss and that the analysis accounted for multiple confounders. Study limitations include its cross-sectional nature and reliance on self-report. The sample may not be representative of the population affected, although the regression model adjusts for multiple demographic factors to aid outcome interpretation. Significant differences were observed between survey modes, with online respondents more likely to report mental health problems and less likely to report physical health problems than telephone respondents. Using multiple response modes and statistically controlling for response mode in regression models may have reduced the response biases commonly observed in health outcomes research (25). Data collection began at the peak of a first wave of COVID-19 cases in Australia and continued through the early stages of re-opening. Longitudinal data from this cohort will track changes in work and employment amongst the study groups, and examine longer-term impacts of mental and physical health as the pandemic unfolds in Australia.

## Data Availability

The data are held at Monash University, Insurance Work and Health Group, School of Public Health and Preventive Medicine. Procedures to access data from this study are available through contacting the lead author. Proposals for collaborative analyses will be considered by the study's investigator team.

## Acknowledgements

We acknowledge the Social Research Centre (SRC) for undertaking telephone interviews.

## Ethics

Approval to conduct the study was provided by Monash University Human Research Ethics Committee.

## References

1. ILO Monitor: COVID-19 and the world of work. Fifth edition Updated estimates and analysis. International Labour Organization. 2020 June 30. url: https://www.ilo.org/wcmsp5/groups/public/---dgreports/---dcomm/documents/briefingnote/wcms_749399.pdf (accessed 11 August 2020).

2. Australian Bureau of Statistics. Labour Force, Australia, July 2020, cat. no. 6202.0; 2020 url: https://www.abs.gov.au/AUSSTATS/abs@.nsf/allprimarymainfeatures/6050C537617B613BCA25836800102753?opendocument (accessed 20 August 2020)

3. Brenner MH. Will There Be an Epidemic of Corollary Illnesses Linked to a COVID-19–Related Recession? American Journal of Public Health. 2020;110, 974–975, 10.2105/AJPH.2020.305724

4. Australian Bureau of Statistics. Household Impacts of COVID-19 Survey, 14-17 April 2020, Australia Canberra; 2020 May 5. Contract No.: 49400DO004_2020.

5. Jane RW Fisher, Thach Duc Tran, Karin Hammargerg, Jayagowri Sastry, Hau Nguyen, Heather Rowe, Sally Popplestone, Ruby Stocker, Claire Stubber and Maggie Kirkman. Mental health of people in Australia in the first month of COVID-19 restrictions: a national survey. Med J Aust [preprint]; 2020. url: https://www.mja.com.au/journal/2020/mental-health-people-australia-first-monthcovid-19-restrictions-national-survey (accessed 20 August 2020).

6. Waddell G, Burton AK. Is work good for your health and well-being? The Stationery Office; 2006 Sep 6.

7. Crowe L, Butterworth P. The role of financial hardship, mastery and social support in the association between employment status and depression: results from an Australian longitudinal cohort study. BMJ open. 2016 May 1;6(5).

8. Zhang SX, Wang Y, Rauch A, Wei F. Health, distress, and life satisfaction of people one-month into COVID-19 outbreak in China. medRxiv. 2020 Jan 1.

9. Taylor MR, Agho KE, Stevens GJ, Raphael B. Factors influencing psychological distress during a disease epidemic: data from Australia's first outbreak of equine influenza. BMC public health. 2008 Dec 1; 8(1): 347.

10. Menec VH, Newall NE, Mackenzie CS, Shooshtari S, Nowicki S. Examining social isolation and loneliness in combination in relation to social support and psychological distress using Canadian Longitudinal Study of Aging (CLSA) data. PLoS One. 2020; 15(3): e0230673.

11. Price RH, Choi JN, Vinokur AD. Links in the chain of adversity following job loss: how financial strain and loss of personal control lead to depression, impaired functioning, and poor health. Journal of occupational health psychology. 2002 Oct;7(4): 302.

12. Morrison, S (Prime Minister of Australia), $ 130 billion JobKeeper payment to keep Australians in a job, media release. 2020 March 30. url: https://www.pm.gov.au/media/130-billion-jobkeeperpayment-keep-australians-job (accessed 30 July 2020)

13. Morrison, S (Prime Minister of Australia) 2020, Supporting Australian workers and business, media release 2020 March 22. url: https://www.pm.gov.au/media/supporting-australian-workers-andbusiness (accessed 30 July 2020)

14. Kessler RC, Andrews G, Colpe LJ, Hiripi E, Mroczek DK, Normand SL, et al Short screening scales to monitor population prevalences and trends in non-specific psychological distress. Psychological medicine. 2002; 32(6): 959–76.

15. Kessler RC, Barker PR, Colpe LJ, Epstein JF, Gfroerer JC, Hiripi E, et al Screening for serious mental illness in the general population. Archives of general psychiatry. 2003; 60(2): 184–9.

16. Ware JE. The SF-12v2TM how to score version 2 of the SF-12® health survey:(with a supplement documenting version 1). Quality metric; 2002.

17. Australian Bureau of Statistics. General Social Survey (GSS): Household Survey Questionnaire. Canberra: Australian Bureau of Statistics; 2014.

18. Koenig HG, Westlund RE, George LK, Hughes DC, Blazer DG, Hybels C. Abbreviating the Duke Social Support Index for use in chronically ill elderly individuals. Psychosomatics. 1993; 34(1): 61–9.

19. Skapinakis P, Weich S, Lewis G, Singleton N, Araya R. Socio-economic position and common mental disorders: Longitudinal study in the general population in the UK. The British Journal of Psychiatry. 2006 Aug;189(2): 109–17.

20. Sheehan LR, Lane TJ, Collie A. The impact of income sources on financial stress in workers’ compensation claimants. Journal of Occupational Rehabilitation. 2020 Feb 27: 1–0.

21. Leigh-Hunt N, Bagguley D, Bash K, Turner V, Turnbull S, Valtorta N, Caan W. An overview of systematic reviews on the public health consequences of social isolation and loneliness. Public Health. 2017 Nov 1; 152: 157–71.

22. Flood M. Mapping loneliness in Australia. Discussion Paper 76. Canberra: Australia Institute; 2005 Feb.

23. Andrews, D (VIC Premier), Statement from the Premier, media release. 2020 June 20. url: https://www.premier.vic.gov.au/statement-premier-71 (accessed 24 August 2020)

24. Australian Bureau of Statistics. Business Indicators, Business Impacts of COVID-19, July 2020, cat. no. 5676. 0.55.003; 2020. url: https://www.abs.gov.au/AUSSTATS/abs@.nsf/mf/5676.0.55.003 (accessed 20 August)

25. Rogler LH, Mroczek DK, Fellows M, Loftus ST. The neglect of response bias in mental health research. The Journal of nervous and mental disease. 2001 Mar 1; 189(3): 182–7.

